# Psychological frailty in older adults: a systematic scoping review

**DOI:** 10.1101/2022.08.01.22278302

**Authors:** Jinlong Zhao, Justina Liu, Stefanos Tyrovolas, Julian Mutz

## Abstract

**Background:** Psychological frailty, along with physical and cognitive frailty, is linked to an increased risk of negative health outcomes among older adults. However, the definition of psychological frailty has received limited attention. A thorough comprehension of the concept of psychological frailty is therefore required.

**Objectives:** To review existing definitions of psychological frailty and to provide a comprehensive overview of the concept of psychological frailty and associated measurements.

**Methods:** This review followed the PRISMA guidelines for scoping reviews and the JBI Manual for Evidence Synthesis. Eligibility criteria were developed based on the Participants-Concept-Context (PCC) framework. We searched CINAHL, Scopus, PubMed, Web of Science, and PsycINFO databases and other sources for relevant studies published between January 2003 to March 2022.

**Results:** The final scoping review included 58 studies. 40 (69%) of these studies provided a definition of psychological frailty and 7 studies provided a novel definition. The other 11 studies focused on components of defining psychological frailty. To better characterize psychological frailty, we propose four groups of components, including mood, cognitive, mental health, and fatigue-associated problems. We identified 28 measuring tools across studies and the Tilburg frailty indicator was the most frequently used (46.6% of studies).

**Conclusions:** Psychological frailty is a complex concept that lacks a consensus definition. It should include both psychological features and physical frailty. Depression and other psychological problems are commonly used to define psychological frailty. This scoping review outlines future research directions to refine the concept of psychological frailty.

## Introduction

Frailty is a clinical syndrome marked by age-associated changes across multiple body systems, weakened physiological functions, and heightened susceptibility to diverse stressors (Dent et al., 2019). Physical frailty emphasizes aging-related physiological changes in the human body. These physiological dysfunctions may contribute to progressive declines in cognitive functioning, also known as cognitive frailty. The concepts of physical and cognitive frailty have been extensively studied in older adults. However, neither concept incorporates the domain of psychological frailty. Like physical and cognitive frailty, psychological frailty results from aging-related decreases in physiological and psychological reserves, contributing to elevated vulnerability to stressors and increased morbidity risks. Frailty as a pre-disability or pre-disease health state necessitates a comprehensive evaluation framework, if health refers to a continuous state of complete physical, psychological, as well as social well-being (WHO, 2006). As such, multiple domains, including psychological aspects, should be integrated into the frailty model.

The importance of comprehensively measuring frailty, including its psychological dimension, has been recognized and highlighted in the literature. Buchman and Bennett (2013) proposed that more studies on frailty that cover psychological and other domains were needed and that such measures may be more sensitive in detecting health issues in older adults. Khezrian, Myint, McNeil, and Murray (2017) suggested that a comprehensive assessment of frailty may better predict adverse health outcomes and help identify potential targets for therapeutic interventions. Researchers have also criticized the absence of a psychological domain in measuring frailty as it may overlook psychosocial issues (De Witte et al., 2013; Hogan, 2018). Most definitions and measurement tools of frailty emphasize physical frailty while omitting psychological features. Psychological frailty, therefore, has been proposed to provide a more comprehensive paradigm of frailty (De Witte et al., 2013).

Only a few studies have examined definitions of psychological frailty or components of psychological frailty (Gobbens, Luijkx, Wijnen-Sponselee, & Schols, 2010a; Shin, Kim, & Choi, 2021). However, there is considerable inconsistency in the conceptualization of psychological frailty. There is also no consensus regarding the assessment of psychological frailty. Indeed, diverse measures (e.g., frailty assessments and mental health scales) have been used to assess psychological frailty, but a specific validated measure is lacking. The absence of a commonly recognized definition of psychological frailty is the likely cause of this.

Current frailty assessments are mostly derived from the cumulative deficit approach developed by Mitnitski and collaborators (2001). This approach was constructed without relevant items clearly indicating psychological frailty. As such, few existing frailty assessments include psychological dimensions. Nevertheless, a few mental health items, such as depression, have been used in some studies to identify psychological plus other frailty domains in the context of frailty indices (Mutz, Choudhury, Zhao, & Dregan, 2022). Incorporating psychological features under the cumulative deficit approach of frailty facilitates a more thorough evaluation of the multiple factors that may lead to an older adult being frail. Moreover, psychological frailty is strongly associated with other frailty domains, such as physical and cognitive frailty (Rietman et al., 2018). A refined definition of psychological frailty could facilitate a deeper understanding of the relationships between different domains of frailty and help disentangle how these may relate to adverse health outcomes. It would also allow for more accurate estimates of psychological frailty prevalence and enable comparisons of findings between studies. However, to date, no study has comprehensively evaluated definitions and components of psychological frailty. Thus, a scoping review of psychological frailty is needed.

### Study objectives

The primary objective was to provide a comprehensive overview of how psychological frailty has been conceptualized in previous studies. The secondary objective was to offer a detailed summary of how psychological frailty has been measured in the available literature.

## Methods

This review adheres to the methodological framework of the Joanna Briggs Institute (JBI) Manual for Evidence Synthesis (Peters et al., 2020) and the PRISMA guidelines extension statement for scoping reviews (PRISMA-ScR) (Tricco et al., 2018).

### Inclusion and exclusion criteria

The inclusion and exclusion criteria were based upon the PCC framework (participants, concept, and context) recommended by the JBI manual (Peters et al., 2020).

#### Types of participants

We included studies involving only or subgroups of older people aged 65 years or over. There were no limitations regarding other demographic characteristics (e.g., sex, ethnicity, or education level).

#### Concept

The core concept, psychological frailty, was examined in this systematic scoping review. Psychological frailty refers to age-related psychological changes involved in the frail brain and mental health problems, and interactions with physical and cognitive frailty. We included literature that provided a definition of the concept of psychological frailty or described any psychological components of frailty. Any literature that only focused on physical frailty without mentioning psychological frailty was excluded.

#### Context

We included studies of older adults living in any setting (e.g., nursing-home, hospitalized, and community-dwelling).

#### Types of included studies

Observational and theoretical studies related to psychological frailty published between January 2003 and March 2022 were considered. Given that the concept of frailty was hypothesized to also include a psychological dimension in 2003 (Markle-Reid & Browne, 2003), articles published before this date were not considered. Only English language publications were included to avoid any misunderstanding during translation.

#### Types of sources

Information sources for this review consisted of electronic databases, grey literature, and books. A literature search was conducted of five electronic databases: Scopus, PubMed, Web of Science (WOS), Cumulative Index to Nursing and Allied Health Literature (CINAHL), and PsycINFO. Additional studies were obtained by screening reference lists of included literature. We also identified grey literature through relevant websites (e.g., databases, Google) and individualized requests to key informants (the author of grey literature). We also set an email alert (PubMed) to receive updates about recently published studies.

### Search strategy

We employed a hybrid syntax of MeSH terms and/or free-text terms to search for relevant literature. The search strategy was based on the three core aspects of this scoping review:

1. Participants: “older adults,” “older people,” “elder,” etc.
2. Concept: “psychological frailty”
3. Context: “geriatric”, “frailty”, etc.

A search strategy was drafted for each database by all authors after consulting an experienced librarian (LN). The full search strategy is presented in **supplementary material 1**.

### Search process

The results of the literature search were imported into a bibliographic software (EndNote version 20.0, Thompson Reuters) and duplicates were deleted. The titles, abstracts, and keywords of identified literature were screened independently by two authors (JZ and JL) to identify potentially relevant studies. The initial findings of this scoping review were compared by both authors to reach an agreement. Full-text articles were then screened separately by two authors for the inclusion and exclusion criteria. Any inconsistencies in the literature selection between the two authors were resolved among all authors (JZ, JL, and ST) to reach a consensus. All reasons for exclusion were documented and are presented in a PRISMA 2020 flowchart (**Figure 1**).

**Figure 1.**
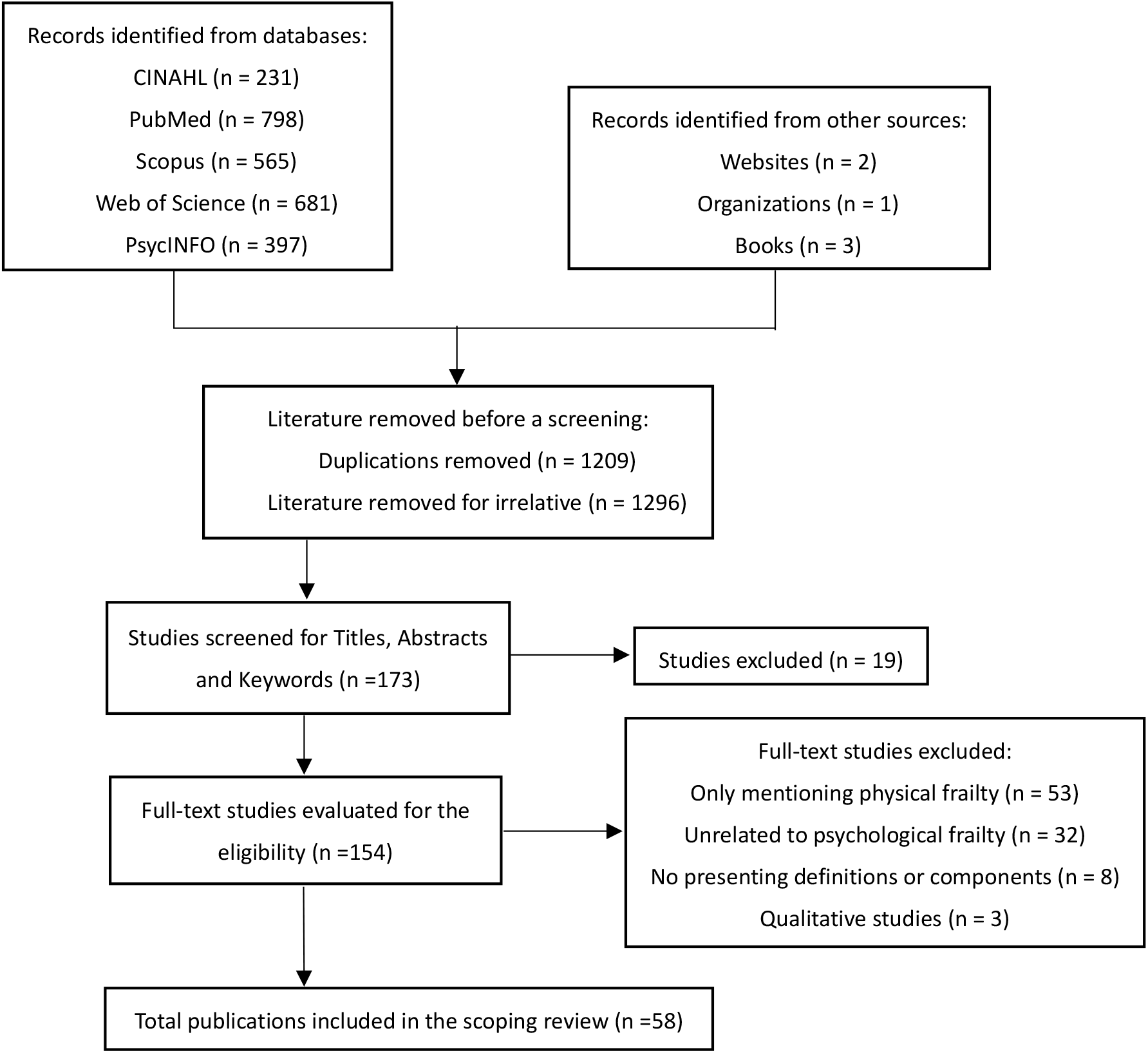
PRISMA flowchart of literature selection

### Data extraction

The characteristics of eligible studies were extracted according to a pre-specified data extraction table which included authors (publication year), countries or regions, research design (sample size), setting, components of psychological frailty, the definition of psychological frailty, and measurements. The details of the extraction were jointly developed by all authors in line with the JBI extraction template (Peters et al., 2020). Data were independently extracted by JZ and JL. Any inconsistencies were resolved by consulting the other authors (TS). If any of the data needed to be clarified, or if key information was missing, the authors of the relevant studies were contacted.

### Strategy for data synthesis

A broad set of definitions and measurements of psychological frailty were entered into the data extraction table for narrative analysis. The definitions and features of psychological frailty were manually grouped and appraised. A more comprehensive definition of psychological frailty or a framework for defining psychological frailty was derived. In the synthesis of the various measurement tools, we coded for each psychological frailty measurement the name of the instrument, the included domains that were assessed, and the number of domains.

## Results

### Characteristics of the publications included

The search yielded 2672 records across the five electronic databases (CINAHL: 231, PubMed: 798, Scopus: 565, Web of Science: 681, and PsycINFO: 397) (**Supplementary material 1**). We identified six additional publications from other sources. After removing duplicates and irrelevant records, 173 publications were further screened. Of these, 19 were excluded after the screening of titles, abstracts, and keywords. The remaining 154 publications were considered for full-text review and 96 publications did not meet our eligibility criteria. 58 publications were included in this scoping review (**Figure 1**).

**Figure 2.**
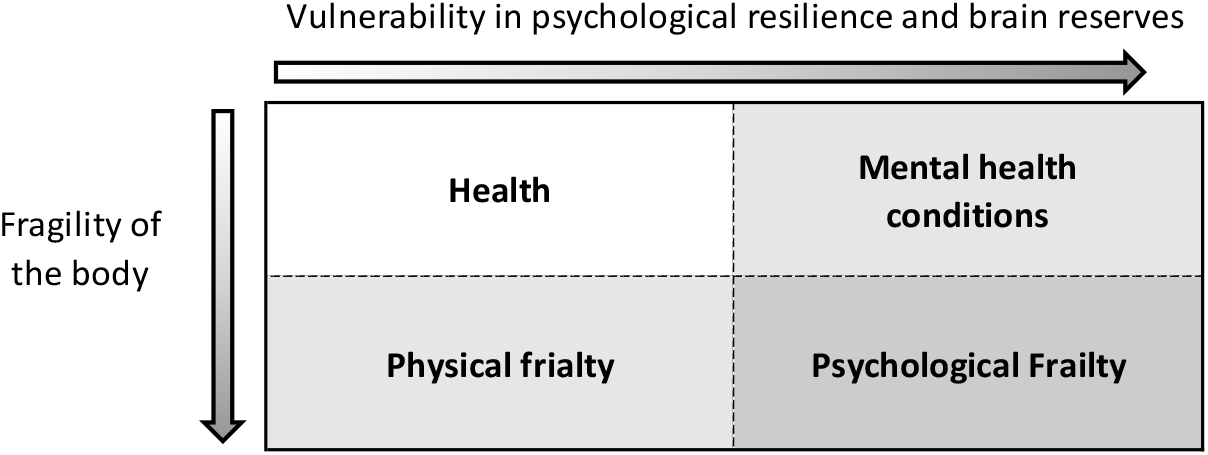
An illustration of the concept of psychological frailty

Of these, thirty-eight (65.5%) studies utilized a cross-sectional design, 16 (27.6%) utilized a longitudinal design, and 4 (6.9%) were theoretical analyses. Over two-thirds of these studies (67.3%) were from European nations and regions, with twenty-one (36.2%) of these conducted in the Netherlands, three in Belgium, two in the United Kingdom, and one in Denmark. The remaining studies included five studies in China (four in mainland China and one in Hong Kong), three in Japan, and two in Canada. A brief description of these studies is presented in **Supplement Table 1**.

### Definitions of psychological frailty

Of the 58 included publications, 40 provided a definition of psychological frailty, with seven focusing on novel definitions of psychological frailty and 33 referencing definitions of other authors. In an attempt to measure psychological frailty, Gobbens and colleagues (2010a) described psychological frailty as a concept that included cognitive, mood, and coping-related features, and they developed a new measurement tool for a comprehensive evaluation of frailty (Gobbens, van Assen, Luijkx, Wijnen-Sponselee, & Schols, 2010b). Twenty-eight of the included studies applied the definition of Gobbens et al. (2010a), indicating that it is frequently used and recognized (**Supplement Table 1**).

De Witte et al. (2013) introduced an operational definition of psychological frailty as the co-existence of loneliness and mood disorders (**Supplement Table 1**). Psychological frailty has also been described as a phenotype of mental frailty that is operationally defined as two or more of the following items: depression, cognitive impairment, low quality of life perception, and low cognitive self-concept (Garre-Olmo, Calvó-Perxas, López-Pousa, de Gracia Blanco, & Vilalta-Franch, 2013). Similarly, Teo et al. (2019) defined mental frailty as meeting one or more of the criteria of low mood, cognitive impairment, and poor self-rated health. As a theoretical construct, Fitten (2015) described psychological frailty as the brain alterations that are deviations from natural aging, but are not necessarily diseases, and contribute to decreased mood or cognitive resilience when confronted with minor stressors, and might induce adverse health results similar to physical frailty. According to Patel et al. (2017), psychological frailty refers to the inherent features of an individual that may predispose an individual to adversity. Rietman et al. (2018) conceptualized psychological frailty as the fulfillment of both criteria for general mental health and depression. Recently, Shimada et al. (2019) defined psychological frailty as the co-existence of depression and physical frailty. Researchers (e.g., Shimada) promoted the importance of considering psychological frailty together with other domains (i.e., physical and cognitive) to capture a comprehensive definition of frailty.

### Components of psychological frailty

All of the included studies provided some information on various components of psychological frailty. We identified five sets of components that were most frequently used to characterize psychological frailty (**Supplement Table 1**).

1. Mood problems are one of the most frequently cited components, including, for example, depression, sadness, and anger (Brehmer-Rinderer, Zeilinger, Radaljevic, & Weber, 2013; Ernsth Bravell et al., 2011). Most researchers have considered mood problems, especially depression, a core component of psychological frailty (Sugie et al., 2022; Ye et al., 2021).
2. Cognitive problems encompass cognitive impairment/deficiency, cognitive symptoms, dementia, poor concentration, memory loss, and related cognitive issues (Garner, Burgess, & Holland, 2020; Gobbens & Andreasen, 2020). Despite some debate, cognitive problems are also one of the most commonly referenced components of psychological frailty.
3. Mental health problems included in the definition of psychological frailty cover a wide spectrum of issues associated with psychological health, for example, anxiety, coping, loneliness, mental disorders, and psychological distress (Gobbens, van Assen, Augustijn, Goumans, & van der Ploeg, 2021a; Hoeyberghs, Verté, Verté, Schols, & De Witte, 2019).
4. Fatigue-associated problems have also been documented in several publications, including fatigue, exhaustion, listlessness, and loss of energy (Schoufour, Mitnitski, Rockwood, Evenhuis, & Echteld, 2013; Shin et al., 2021). These problems likely overlap with the frailty phenotype model (Fried et al., 2001). Fatigue-associated problems have been highlighted as a key component of psychological frailty (Shimada et al., 2019).

### Defining criteria and measurements of psychological frailty

Overall, 28 measurement tools were employed to assess psychological frailty across 54 studies (**Supplement Table 1**). Of these, ten studies provided their own measurement tools of psychological frailty or a psychological domain of frailty. The most frequently used measurement tool of psychological frailty was the Tilburg Frailty Indicator (TFI). It is a self-report scale developed by Gobbens et al. (2010b). TFI covers physical (eight questions), social (three questions), and psychological (four questions) domains. The psychological domain is assessed using four yes/no questions on cognition, anxiety, depression, and coping. From these, a score of psychological frailty that ranges from zero to four can be derived. A higher value represents a more severe level of psychological frailty. Another assessment tool is the Groningen Frailty Indicator (GFI). It is a common screening tool of frailty, composed of four domains (fifteen questions). The psychological domain is assessed using two questions on sadness and anxiousness. Individuals with a total score of at least one on this dimension are considered psychologically frailty (Ament, Vugt, Verhey, & Kempen, 2014; Steverink, Slaets, Schuurmans, & Lis, 2001).

Furthermore, various mental health assessment tools have been used to measure psychological frailty. Shimada et al. (2019) utilized the 15-item geriatric depression scale (GDS-15) and Fried’s frailty phenotype to screen for psychological frailty. These authors defined psychological frailty as coexistence of depression (GDS-15 score of at least four or five) and physical frailty (meeting at least three criteria of the frailty phenotype). Nishida, Yamabe, and Honda (2020) only measured depressive mood (defined as a score of at least two on the depression domain of the frailty checklist) to represent psychological frailty. The Center for Epidemiologic Studies Depression Scale (CES-D) and the five-item Mental Health Inventory (MHI-5) were also used to assess psychological frailty, defined as screening positive for on both MHI-5 and CES-D (Rietman et al., 2018). Details of other measurement tools of psychological frailty are presented in **Supplement Table 1**.

## Discussion

### Existing definitions, components, and measurements

This review aimed to provide an overview of existing definitions and measurement of psychological frailty. Despite increasing empirical studies examining psychological frailty in recent years, few studies have explored its definition. Most studies did not clearly articulate a theoretical definition or framework of psychological frailty (Gobbens & Andreasen, 2020; Venturini et al., 2021). Some studies have defined psychological frailty according to a single or a small number of mental health items, for example, depression (Sugie et al., 2022). There was little to no consideration of the difference between mental disorders and psychological frailty and what “frailty” implied in specific psychological domains. Psychological frailty, as a subtype of frailty, should be more closely linked to the risks associated with mental disorders (**Figure** 2). Otherwise, the concept of psychological frailty would extensively overlap with mental disorders. Frailty in the psychological context is multifactorial and incorporates not only features of psychological functioning but also physical frailty-related elements, such as fatigue and exhaustion.

Psychological frailty may result from aging-related deficits in psychological resilience and brain reserves. Psychological resilience refers to the capacity of an individual to cope in the face of stressors and adversity, allowing individuals to successfully maintain their physical and mental health (Afek et al., 2021; Fletcher & Sarkar, 2013). Brain reserve is defined as the ability of an individual to withstand aging-associated and pathological changes to the brain, also known as resilience of the brain (Fratiglioni & Wang, 2007). Aging-associated declines in psychological resilience and brain reserve indicate the progressive impact of aging on the brain and the emergence of psychological frailty. As such, psychological frailty may be described as the decreased state of psychological resilience and brain reserve caused by various physiological aging processes in the brain. This state is potentially reversible, analogous to cognitive frailty (Bémeur & Rose, 2020). However, more evidence is needed to validate this hypothesis.

While several studies have explored how psychological frailty may be defined, we discovered that the various definitions lacked consistency and consensus. For example, one study defined psychological frailty as the co-existence of depression and physical frailty (Shimada et al., 2019). In contrast to this definition, other studies defined psychological frailty without including physical frailty as a component (Patel, 2017; Rietman et al., 2018). Cognitive components were incorporated in the definition of psychological frailty in some studies (Fitten, 2015; Gobbens et al., 2010b), while they were not considered in other definitions (Teo et al., 2019). A possible explanation for this inconsistency is the lack of agreement on the theoretical foundation of psychological frailty. This concern also applies to the general concept of frailty which lacks a clear and agreed-upon theoretical foundation (Bergman et al., 2007). Another potential explanation is the lack of effort to obtain a consensus definition of psychological frailty amongst researchers. Although more studies have attempted to incorporate psychological domains in research on frailty, few have specifically focused on conceptualizing psychological frailty. Our scoping review showed that there were only seven studies over the past two decades that provided novel definitions of psychological frailty.

Several other issues regarding the conceptualization of psychological frailty deserve further attention. For example, one issue is how to select diverse psychological functions to define psychological frailty. We found that psychological functions incorporated into definitions of psychological frailty overlapped in numerous studies (Shimada et al., 2019; Venturini et al., 2021; Ye et al., 2021). However, the lack of consistent criteria for selecting psychological functions makes it difficult to reach a consensus definition. There was little evidence to establish associations between individual psychological functions and the overall concept of psychological frailty. In our review, numerous individual components reported across studies were classified into five groups, including mood, cognitive, mental health, fatigue-associated, and other problems. Although this classification may be imperfect, it serves as an attempt to systematically arrange these components into multiple categories based on similarity. Empirical studies are needed to validate these five categories. Our scoping review also revealed that, whereas physical and cognitive problems (physical and cognitive frailty) are frequently incorporated components of psychological frailty, overlap and distinctions between these domains within a comprehensive framework of frailty have not been thoroughly examined (van Oostrom et al., 2017).

Another issue to be noted is that more accurate measurements of psychological frailty are needed. Our review demonstrated that most measurement tools were designed to assess one or several psychological functions, mental disorders, or comprehensive frailty, while tools built specifically to assess psychological frailty were not identified. Nevertheless, psychological frailty may be assessed as part of comprehensive frailty assessments, such as the Groningen Frailty Index and Tilburg Frailty Indicator.

As psychological frailty may co-exist with other frailty domains, both the deficit accumulation model and Fried’s frailty phenotype together with psychological items have frequently been used to measure psychological frailty. The deficit accumulation model of frailty (Rockwood & Mitnitski, 2011) may be more suitable to assess psychological frailty and was also used more widely across studies, compared to the frailty phenotype. The deficit accumulation model can incorporate any type of health deficit, also including psychological deficits. As such, an increasing number of studies have adopted this approach to create measurement tools of psychological frailty. For example, Kwan et al. (2015) constructed a comprehensive frailty model and assessed psychological domains using items on positive and negative psychological well-being according to the cumulative deficit approach. Whether these measures are valid to assess psychological frailty remains unknown as a clear definition of psychological frailty is lacking and there is no gold standard to compare these different measurement tools. As noted above, the reliability and validity of several measurement tools (as tools of psychological frailty) in the included studies have not been assessed (Huang & Lam, 2021).

### Implications of future research

A comprehensive and consensus definition of psychological frailty is needed. First, a key strategy should be established to form a firm theoretical framework of psychological frailty through group discussions among experts from multiple research areas. These individuals should have expertise in frailty, mental health, geriatrics, nursing, and related fields. A consensus statement should be developed to address the question “what is psychological frailty?”. This would likely incorporate various psychological functions (e.g., perception, attention, reasoning, mood, language, and memory) and multiple items of physical frailty reflecting the complexities of psychological frailty and characterizing psychological frailty as a flexible and multifaceted concept. Second, the operational definition of psychological frailty based on this theoretical framework should be specific. An appropriate operational definition could provide a foundation for designing successful and valid measurement tools to assess psychological frailty.

Future research also needs to clarify the boundaries between psychological frailty and other domains of frailty, including physical and cognitive frailty. This could help identify possible overlap and differences between the definitions of these three domains of frailty. Indeed, these domains may be inherently interconnected and result from normal and pathological aging (Garre-Olmo et al., 2013). Future research should also develop a specific scale to assess psychological frailty. This effort is important because there are no specific and valid measurement tools of psychological frailty. Ideally, both subjective and objective measures (including self-report data and biomarkers) should be included in the measurement of psychological frailty. Items from existing gold standard measurements such as Fried’s frailty phenotype and the DSM-5 criteria should be incorporated where possible. The psychometric characteristics of such measurement tools of psychological frailty should also be systematically investigated.

### Limitations of the study

A potential limitation of this review might be the search strategy. The focus of our review was the concept of psychological frailty and we did not include similar concepts, such as mind, or emotional frailty. These similar concepts were not included because they are rarely investigated and applied in existing research. Another limitation is that the psychometric properties of different measurement tools used to identify psychological frailty were not explored in this review.

## Conclusions

Psychological frailty is a complex concept with no consensus definition. As a multifactorial concept, psychological frailty should include both mental features and physical frailty. Depression and other problems are frequently used as core components of psychological frailty. Also, this study has presented several future research directions for better conceptualizing psychological frailty. A more precise definition and improved understanding of the concept of psychological frailty might encourage its use in clinical settings and might contribute to an improved description of frailty at the psychological or neural level.

## Data Availability

All data reported here were sourced from existing publications.

## Ethics

Ethical approval was not needed because this systematic scoping review did not directly involve any participants and only relied on published documents for analysis.

## Acknowledgments

We thank our librarian (Ms Lydia Ngai; LN) of the Faculty of Health and Social Sciences at the Hong Kong Polytechnic University who provided specialized advice regarding the literature search strategy.

## Author contribution

JZ designed the research, searched the literature, selected studies for inclusion, analyzed the results, drafted this manuscript, and revised this manuscript for final submission. JL contributed to the research design, developed inclusion criteria, and critically reviewed this manuscript. ST and JM participated in discussions and reviewed some versions of this manuscript. All authors read and approved the final manuscript.

## Data availability

All data reported here were sourced from existing publications.

## Supplementary materials

Supplementary material is available online.

