## Supplementary Materials for "Psychological frailty in older adults: a systematic scoping review"

### 1. **The complete search strategy** (5 databases)

The following search strategy was used for the literature search.
Years of publication covered: 2003-2022.
Limitations: humans, English language.
Search date: 11 March 2022.

CINAHL Complete (via EBSCO Host) Search:

| | | |
|---|---|---|
| #1 | Frailty* [TX] OR Frail* [TX] OR “Frailty Syndrome” [MH] OR Physical frailty [TX] | 43,063 |
| #2 | Psychological frailty [TX] | 256 |
| #3 | #1 AND #2 | 256 |
| #4 | (elder* OR geriatric* OR “old age” OR “oldest old” OR “older old” OR “later life” OR ageing OR aging OR “old adults” OR “older adults” OR “oldest adults” OR “old population” OR “Older population” OR “oldest population” OR “old individuals” OR “older individuals” OR “oldest individuals” OR “old patients” OR “older patients” OR “oldest patients” OR “old persons” OR “older persons” OR “oldest persons” OR “old people” OR “older people” OR “oldest people” OR “aged adults” OR “aged population” OR “aged individuals” OR “aged patients” OR “aged persons” OR “aged people”) [TX] | 670,818 |
| #5 | “Frail Elderly” [MH] | 8,300 |
| #6 | #4 OR #5 | 670,818 |
| #7 | #3 AND #6 | 239 |
| #8 | Limit to “English” and “published year after 2003” | |
| #9 | #7 AND #8 | 231 |

PubMed Search:

| | | |
|---|---|---|
| #1 | “Frailty” [All Fields] OR “Frail” [All Fields] OR “Physical frailty” [All Fields] | 34,974 |
| #2 | “Psychological frailty” [All Fields] | 982 |
| #3 | #1 AND #2 | 977 |
| #4 | (elder* OR geriatric* OR “old age” OR “oldest old” OR “older old” OR “later life” OR ageing OR aging OR “old adults” OR “older adults” OR “oldest adults” OR “old population” OR “Older population” OR “oldest population” OR “old individuals” OR “older individuals” OR “oldest individuals” OR “old patients” OR “older patients” OR “oldest patients” OR “old persons” OR “older persons” OR “oldest persons” OR “old people” OR “older people” OR “oldest people” OR “aged | 1,044,510 |

| | | |
|---|---|---|
| | adults” OR “aged population” OR “aged individuals” OR “aged patients” OR “aged persons” OR “aged people”) | |
| #5 | “Aged” [MeSH] OR “Frail Elderly” [MeSH] | 3,380,462 |
| #6 | #4 OR #5 | 3,948,917 |
| #7 | #3 AND #6 | 880 |
| #8 | Limit to “English” and “published year after 2003” | |
| #9 | #7 AND #8 | 798 |

Scopus Search:

(TITLE-ABS-KEY (frailty*) AND TITLE-ABS-KEY (frail*) AND TITLE-ABS-KEY (physical frailty) AND TITLE-ABS-KEY (psychological frailty)) AND (TITLE-ABS-KEY (elder* OR geriatric* OR “old age” OR “oldest old” OR “older old” OR “later life” OR ageing OR aging OR “old adults” OR “older adults” OR “oldest adults” OR “old population” OR “Older population” OR “oldest population” OR “old individuals” OR “older individuals” OR “oldest individuals” OR “old patients” OR “older patients” OR “oldest patients” OR “old persons” OR “older persons” OR “oldest persons” OR “old people” OR “older people” OR “oldest people” OR “aged adults” OR “aged population” OR “aged individuals” OR “aged patients” OR “aged persons” OR “aged people”)) AND (SUBJAREA (nurs OR medi OR heal OR mult OR psyc OR soci)) AND (LANGUAGE (English)) AND (PUBYEAR AFT 2003)

=565 articles

Web of Science Search:

| | | |
|---|---|---|
| #1 | TS= (frailty*) OR TS= (frail*) OR TS= (physical frailty) | 39,158 |
| #2 | TS= (psychological frailty) | 810 |
| #3 | #1 AND #2 | 810 |
| #4 | TS= (elder* OR geriatric* OR “old age” OR “oldest old” OR “older old” OR “later life” OR ageing OR aging OR “old adults” OR “older adults” OR “oldest adults” OR “old population” OR “Older population” OR “oldest population” OR “old individuals” OR “older individuals” OR “oldest individuals” OR “old patients” OR “older patients” OR “oldest patients” OR “old persons” OR “older persons” OR “oldest persons” OR “old people” OR “older people” OR “oldest people” OR “aged adults” OR “aged population” OR “aged individuals” OR “aged patients” OR “aged persons” OR “aged people”) | 3,894,104 |
| #5 | #3 AND #4 | 724 |
| #6 | Limit to “English” and “published year after 2003” | |
| #7 | #6 AND #7 | 681 |

PsycINFO (via ProQuest) Search:

| | | |
|---|---|---|
| #1 | Frailty* [Anywhere] OR Frail* [Anywhere] OR Physical frailty [Anywhere] | 6,495 |
| #2 | Psychological frailty [Anywhere] | 663 |
| #3 | #1 AND #2 | 663 |
| #4 | (elder* OR geriatric* OR “old age” OR “oldest old” OR “older old” OR “later life” OR ageing OR aging OR “old adults” OR “older adults” OR “oldest adults” OR “old population” OR “Older population” OR “oldest population” OR “old individuals” OR “older individuals” OR “oldest individuals” OR “old patients” OR “older patients” OR “oldest patients” OR “old persons” OR “older persons” OR “oldest persons” OR “old people” OR “older people” OR “oldest people” OR “aged adults” OR “aged population” OR “aged individuals” OR “aged patients” OR “aged persons” OR “aged people”) [TIAB] | 183,376 |
| #5 | #3 AND #4 | 456 |
| #6 | Limit to “English” and “published year after 2003” | |
| #7 | # 5 AND #6 | 397 |

**Supplement Table 1.**

**Supplement Table 1.** Characteristics of included studies

| Authors (year) | Countries or regions | Research design (sample size) | Setting | Components of psychological frailty | Definition of psychological frailty | Measurements | |
|---|---|---|---|---|---|---|---|
| | | | | | | Instruments | Number of components |
| Puts et al. (2005) | Netherlands | Longitudinal (n=1,152) | Community | Sense of mastery and depression | n/a | CES-D<br>SOMS-7 | 2 |
| Rockwood et al. (2005) | Canada | Prospective cohort (n=2,305) | Community or institution | Mood problems, depression, restlessness, memory, cognitive symptoms, paranoid features, clouding or delirium, and general mental functioning | n/a | CSHA-FI | 8 |
| Rolfson et al. (2006) | Canada | Cross-sectional (n=158) | Inpatient and outpatient | Mood | n/a | EFS | 1 |
| Gu et al. (2009) | China | Longitudinal (n=15,919) | Any settings | Symptom of psychological distress (loneliness, usefulness, and fearfulness) | n/a | FI-39 | 3 |
| Gobbens et al. (2010a) | n/a | Theoretical (n/a) | n/a | Cognition, emotion, and coping | Psychological variables related to frailty (Psychological domain of frailty) | n/a | n/a |
| Gobbens et al. (2010b) | Netherlands | Cross-sectional (n=245;234) | Community | Coping, anxiety, depressive symptoms, and cognition | Definition of Gobbens et al. (2010a) | TFI<br>MMSE<br>GES-D<br>HANS-A<br>SOMS-5 | 4 |
| Gobbens et al. (2010c) | Netherlands | Cross-sectional (n=484) | Community | Coping, anxiety, depressive symptoms, and cognition | Definition of Gobbens et al. (2010a) | TFI | 4 |

| | | | | | | | |
|---|---|---|---|---|---|---|---|
| Kamaruzzaman et al. (2010) | United Kingdom | Cohort (n=4,286) | Community | Psychological problems (Memory, anxiety, and depression) | n/a | BFI | 3 |
| Ernsth Bravell et al. (2011) | Sweden | Cross-sectional (n=315) | Community | Anxiety/insecurity, sadness/gloom, cognitive deficiency, and behaviors that are hard to handle/manage | n/a | A scale of psychological/cognitive symptoms in SNAC study | 4 |
| Gobbens et al. (2012a) | Netherlands | Cross-sectional (n=213) | Community | Coping, anxiety, depressive symptoms, and cognition | Definition of Gobbens et al. (2010a) | TFI | 4 |
| Gobbens et al. (2012b) | Netherlands | Longitudinal (n=484;336;266) | Community | Coping, anxiety, depressive symptoms, and cognition | Definition of Gobbens et al. (2010a) | TFI | 4 |
| Gobbens et al. (2012c) | Netherlands | Longitudinal (n=245;179;141) | Community | Coping, anxiety, depressive symptoms, and cognition | Definition of Gobbens et al. (2010a) | TFI | 4 |
| Bielderman et al. (2013) | Netherlands | Cross-sectional (n=1,508) | Community | Psychosocial functioning (Sadness and anxiousness) | n/a | GFI | 2 |
| Brehmer-Rinderer et al. (2013) | Austria | Cross-sectional (n=147) | Clinical setting | Fear of falling, memory, exhaustion, anxiety, sadness, anger and other psychological changes | n/a | VFQ-ID-R | 7 |
| De Witte et al. (2013) | Belgium | Cross-sectional (n=33,629) | Community | Mood disorders and loneliness | A combination of mood disorders and loneliness | CFAI | 2 |
| Garre-Olmo et al. (2013) | Spain | Prospective cohort (n=875) | Community | Suspected cognitive impairment, suspected depression, low cognitive self-concept, and low quality of life perception | Mental frailty phenotype (MFP) operationally defined as two or more MFP criteria | MMSE<br>GDS-5<br>SAPS<br>VAS | 4 |
| Schoufour et al. (2013) | Netherlands | Cross-sectional (n=1,050) | Care organizations | Fatigue, listlessness, and panic attacks | n/a | Dutch version of ADESS | 3 |

| | | | | | | | |
|---|---|---|---|---|---|---|---|
| Ament et al. (2014) | Netherlands | Longitudinal (n=475) | Community | Downheartedness and anxiety | n/a | GFI | 2 |
| Andreasen et al. (2014) | Denmark | Cross-sectional (n=34) | Community center and acute medical ward | Coping, anxiety, depressive symptoms, and cognition | Definition of Gobbens et al. (2010a) | Danish version of TFI | 4 |
| Gobbens et al. (2014) | Netherlands | Longitudinal (n=261;196) | Community | Coping, anxiety, depressive symptoms, and cognition | Definition of Gobbens et al. (2010a) | TFI | 4 |
| Coelho et al. (2015) | Portugal | Cross-sectional (n=252) | Community | Coping, anxiety, depressive symptoms, and cognition | Definition of Gobbens et al. (2010a) | TFI | 4 |
| Fitten (2015) | n/a | Theoretical (n/a) | n/a | Cognitive, mood, and motivational components | Brain alterations that are beyond natural aging, but are not necessarily diseases, and contribute to decreased mood or cognitive resilience when confronted with minor stressors, and might induce adverse health outcomes similar to physical frailty | n/a | 3 |
| Gobbens et al. (2015) | Netherlands | Cross-sectional (n=221) | Assisted living facilities | Coping, anxiety, depressive symptoms, and cognition | Definition of Gobbens et al. (2010a) | TFI | 4 |
| Kwan et al. (2015) | Hong Kong | Cross-sectional (n=124) | Community | FI-32: Psychocognitive well-being (psychiatric disorders, dementia and hopelessness); FI-32 Plus: (A) positive psychological well-being (4 items) and (B) negative psychological well-being (3 items) | n/a | FI-32<br>FI-32 Plus | 10 |
| Peters et al. (2015) | Netherlands | Cohort (n=5,712) | Community | Psychosocial functioning | n/a | GFI | 5 |

| | | | | | | | |
|---|---|---|---|---|---|---|---|
| Roppolo et al. (2015) | Italy | Cross-sectional (n=267) | Community | Coping, anxiety, depressive symptoms, and cognition | Definition of Gobbens et al. (2010a) | Italian version of TFI | 4 |
| Freitag et al. (2016) | Germany | Cross-sectional (n=210) | Community | Coping, anxiety, depressive symptoms, and cognition | Definition of Gobbens et al. (2010a) | German version of TFI | 4 |
| Gobbens et al. (2016) | Netherlands | Cross-sectional (n=610) | Community | Coping, anxiety, depressive symptoms, and cognition | Definition of Gobbens et al. (2010a) | TFI | 4 |
| Uchmanowicz et al. (2016) | Poland | Cross-sectional (n=212) | Community | Coping, anxiety, depressive symptoms, and cognition | Definition of Gobbens et al. (2010a) | Polish version of TFI | 4 |
| Coelho et al. (2017) | Portugal | Cross-sectional (n=252) | Community | Coping, anxiety, depressive symptoms, and cognition | Definition of Gobbens et al. (2010a) | TFI | 4 |
| Dong et al. (2017) | China | Cross-sectional (n=917) | Community | Coping, anxiety, depressive symptoms, and cognition | Definition of Gobbens et al. (2010a) | Chinese version of TFI | 4 |
| Gobbens et al. (2017) | Netherlands | Cross-sectional (n=671) | Community | Coping, anxiety, depressive symptoms, and cognition | Definition of Gobbens et al. (2010a) | TFI | 4 |
| Patel et al. (2017) | n/a | Theoretical (n/a) | n/a | Bereavement, low mood, lack of motivation, and labile emotions | Psychological frailty denotes the inherent traits that may predispose a person to adversity | n/a | n/a |
| van Oostrom et al. (2017) | Netherlands | Cross-sectional (n=4,019) | Community | Depression and poor mental health | Adapted from Gobbens et al. (2010a) | CES-D<br>MHI-5 | 2 |
| Hogan (2018) | n/a | Theoretical (n/a) | n/a | Cognition, depression, and psychological attributes (e.g., positive affect) | n/a | n/a | n/a |

| | | | | | | | |
|---|---|---|---|---|---|---|---|
| Ma et al. (2018) | China | Cross-sectional (n=5,844) | Community | Poor mental health | n/a | CGA-FI<br>CES-D | 1 |
| Rietman et al. (2018) | Netherlands | Cross-sectional (n=4,019) | Community | Depression and mental health problems | Psychological frailty defined as meeting both criteria for poor general mental health and depression | CES-D<br>MHI-5 | 2 |
| Vrotsou et al. (2018) | Spain | Prospective cohort (n=865) | Community | Coping, anxiety, depression, and cognition | Definition of Gobbens et al. (2010a) | Spanish version of TFI | 4 |
| Hoeybergs et al. (2019) | Belgium | Cross-sectional (n=16,872) | Community | Mood disorders and loneliness | Definition of De Witte et al. (2013) | CFAI | 2 |
| Salem et al. (2019) | Los Angeles and Pomona (United States) | Cross-sectional (n=130) | Formerly incarcerated and homeless | Coping, anxiety, depression, and cognition | Definition of Gobbens et al. (2010a) | TFI | 4 |
| Shimada et al. (2019) | Japan | National cohort (n= 4,126) | Any settings | Physical frailty and depression | The coexistence of depression and physical frailty | FFP<br>GDS-15 | 2 |
| Teo et al. (2019) | Singapore | Cross-sectional (n=2,387) | Community | Poor self-rated health, low mood, and cognitive impairment | Mental Frailty (MF) defined as meeting one or more of the MF criteria | MMSE<br>GDS-15<br>SF-12 | 3 |
| Verver et al. (2019) | Netherlands | Cross-sectional (n=1,768) | Community | Coping, anxiety, depression, and cognition | Definition of Gobbens et al. (2010a) | TFI | 4 |

| | | | | | | | |
|---|---|---|---|---|---|---|---|
| Zhang et al. (2019) | United Kingdom, Greece, Croatia, Netherlands, and Spain | Cross-sectional (n=2,167) | Community | Coping, anxiety, depression, and cognition | Definition of Gobbens et al. (2010a) | TFI | 4 |
| Alqahtani et al. (2020) | Saudi Arabia | Cross-sectional (n=84) | Community | Coping, anxiety, depression, and cognition | Definition of Gobbens et al. (2010a) | Arabic (Saudi) version of TFI | 4 |
| Garner et al. (2020) | United Kingdom | Longitudinal (n=351) | Community and retirement village | 17 psychological markers (depressed mood, dementia, psychiatric disorders, etc.) | n/a | COM-FI | 17 |
| Gobbens et al. (2020) | Netherlands | Prospective cohort (n=1,328) | Community | Cognition, depression, anxiety, and coping | Definition of Gobbens et al. (2010a) | TFI | 4 |
| Nishida et al. (2020) | Japan | Cross-sectional (n=3,475) | Community | Depression | n/a | FC | 1 |
| Ožić et al. (2020) | Rijeka (Croatia) | Prospective interventional (n=410) | Community | Cognition, anxiety, depression, and coping | Definition of Gobbens et al. (2010a) | TFI | 4 |
| Zhang et al. (2020) | China | Cross-sectional (n=5,341) | Community | Six items on mental state (poor concentration, sadness or depression, etc.) | n/a | FI-33 | 6 |
| Gobbens et al. (2021a) | Netherlands | Longitudinal (n=479) | Community | Cognition, anxiety, depression, and coping | Definition of Gobbens et al. (2010a) | TFI | 4 |
| Gobbens et al. (2021b) | Netherlands | Longitudinal (n=479) | Community | Cognition, anxiety, depression, and coping | Definition of Gobbens et al. (2010a) | TFI | 4 |
| Shin et al. (2021) | Korea | Cross-sectional (n=2,923) | Community | Fatigue and loss of energy | n/a | KFS | 2 |

| | | | | | | | |
|---|---|---|---|---|---|---|---|
| Van der Elst et al. (2021) | Belgium | Cross-sectional (n=12,659) | Community | Mood disorders and loneliness | Definition of De Witte et al. (2013) | CFAI | 2 |
| Venturini et al. (2021) | Brazil | Cross-sectional (n=3,569) | Community | Memory problems, depressive symptoms, sadness, and concern about falls | Adapted from Gobbens et al. (2010a) | GDS-15 Brazil<br>FES-I Brazil | 4 |
| Ye et al. (2021) | United Kingdom, Greece, Croatia, Netherlands, and Spain | Cross-sectional (n=2,325) | Community | Cognition, anxiety, depression, and coping | Definition of Gobbens et al. (2010a) | TFI | 4 |
| Sugie et al. (2022) | Japan | Cross-sectional (n=577) | Village | Depression | n/a | GDS-15 | 1 |
| van Assen et al. (2022) | Netherlands | Cross-sectional (n=45,336) | Community | Cognition, anxiety, depression, and coping | Definition of Gobbens et al. (2010a) | TFI | 4 |

Notes: n/a = Not Applicable or Not Available; ADESS = Anxiety, Depression And Mood Scale; BFI = British Frailty Index; GDS-5 = 5-item Geriatric Depression Scale; GDS-15 = 15-item Geriatric Depression Scale; CES-D = Center for Epidemiologic Studies Depression Scale 20 items version; CFAI = Comprehensive Frailty Assessment Instrument; CGA-FI = Comprehensive Geriatric Assessment-Frailty Index; COM-FI = Community-Oriented Frailty Index; CSHA = Canadian Study of Health and Aging; EFS = Edmonton Frail Scale; FC = Frailty Checklist (Yamada et al., 2012); FES-I Brazil = Brazilian version of the Falls Efficacy Scale–International; FFP = Fried's Frailty Phenotype; FI = Frailty Index; GFI = Groningen Frailty Indicator; HADS-A = Hospital Anxiety and Depression Scale-anxiety subscale; HRQOL = Health-related Quality of Life; KFS = Korean Frailty Scale; MHI-5 = Mental Health Inventory 5 items version; MMSE = Mini-Mental State Examination; QOL = Quality of Life; SAPS = Subjective Aging Perception Scale; SF-12 = 12-item Short Form Survey; SNAC = Swedish National Study on Aging and Care; SOMS-5 = 5-item version of the Sense of Mastery Scale; SOMS-7 = 7-item version of the Sense of Mastery Scale; TFI = Tilburg Frailty Indicator; VAS = Visual Analogue Scale; VFQ-ID-R = Vienna Frailty Questionnaire-Intellectual Disabilities – Revised
